# Understanding widening socio-economic inequities in cardiovascular disease mortality in the Netherlands: A mixed methods protocol

**DOI:** 10.64898/2026.10.23.26366312

**Authors:** Kristina Thompson

**Affiliations:** Health & Society, Wageningen University & Research, the Netherlands

**Keywords:** socio-economic inequity, health, mixed methods, group model building, microsimulation

## Abstract

**Background:** In the Netherlands, there is a substantial social gradient in mortality: individuals with higher socio-economic status are more likely to live longer lives than those with lower socio-economic status. There is evidence that this social gradient has widened since the mid-twentieth century, although why this has occurred is poorly understood.

Cardiovascular disease (CVD) provides a particularly useful case study for understanding the mechanisms underlying this widening gradient. While CVD was historically the leading cause of death in the Netherlands, improvements in prevention and treatment have decreased CVD mortality, even as socio-economic inequalities in CVD mortality have increased. In this project, I aim to identify and test the mechanisms driving widening socio-economic inequities in CVD mortality over the last 75 years.

**Methods and analysis:** This project uses mixed-methods. First, in participatory workshops, key stakeholders, e.g. policymakers, healthcare professionals, patient representatives, and individuals with lived experience of poverty, will co-create a conceptual model outlining the explanations linking socio-economic status and CVD in the Netherlands. Second, the conceptual model will provide the structure for the microsimulation, with the former defining which mechanisms will be represented in the latter. The microsimulation will maintain a common model structure across time, although population characteristics and individual trajectories will change to reflect successive twenty-year periods from the 1950s until today. The microsimulation will be validated by comparing simulated outcomes with historical data. Counterfactual scenarios will then be used to estimate the relative contribution of different mechanisms to the widening of socio-economic inequalities in CVD mortality.

**Ethics and dissemination:** This project has been approved by the Social Sciences Research Ethics Committee of Wageningen University (approval number 2026-203). Participants will provide informed consent prior to participation. The microsimulation code will be made openly available, and findings will be disseminated through open-access publications, conference presentations, and a public summary co-designed with stakeholders.

**Article summary Strengths and limitations:**

- Integrating qualitative stakeholder input is expected to improve the face validity of the microsimulation.
- Parameterizing the same microsimulation framework across successive 20-year historical periods allows changes in the contribution of different mechanisms to widening socio-economic inequities in health to be quantified.
- Historical data are more limited than contemporary linked data, meaning that some model parameters may need to be estimated using proxy measures or assumptions.
- Mechanisms identified through participatory sessions may differ from those identified as important in the existing literature

## **1.** Introduction

### Background

Today in the Netherlands, there are substantial socio-economic differences in health and mortality: people with higher socio-economic status (SES) tend to live healthier, longer lives than those with lower SES.(1) Dutch people in the top 10% of the income distribution live an average of eight years longer than those in the bottom 10%.(2) Such differences in health are inequities, rather than simply inequalities, because these differences are avoidable, unjust and systemic. Socio-economic inequities in mortality are also at odds with the principles of the Dutch modern welfare state, and are extremely costly for the Dutch healthcare system.(3,4) Healthcare expenditure was estimated to be between 50% and 150% higher for people in the lowest SES groups, compared to those in the highest SES groups.(3) Reducing socio-economic inequities in health is therefore a longstanding public health priority of the Dutch government.(5)

However, these efforts have partly failed. Although lifespan and the number of years lived in good health have increased in recent decades, several measures of socio-economic inequities have either persisted, or, as in the case of mortality, have appeared to widen. In 2018, the Netherlands’ Scientific Council for Government Policy (WRR) found that policies aimed at reducing differences in health between high and low socio-economic groups often resulted in health improvements for *higher* socio-economic groups.(6)

Further, there is evidence that inequities in mortality have increased since the mid-twentieth century. In a review of the literature from the 1950s to 1980s, Mackenbach(7) found that studies in the 1950s and 1960s infrequently found an association between socio-economic status and mortality, while those from the 1970s onward found clear and widening associations. Exploring the social gradient in mortality among adult men dying between 1870 and 1984, Thompson & van Ophem(8) similarly found the first evidence of a social gradient in mortality born in the early twentieth century (and generally died from the mid-twentieth century onward). In other words, the social gradient in mortality, especially among adults, appears to be a modern phenomenon that emerged in the middle of the twentieth century. However, why exactly the social gradient has widened over this period is an open question.(4)

Answering this question is a critical first step to ultimately reducing differences between high and low socio-economic groups in the Netherlands, and to improving population health overall.(9,10) The Netherlands’ Council for Public Health and Society(11) wrote that, in order to be able to address health inequities, “*It is worth remembering that we are currently seeing the results of what happened years ago rather than what is happening now. It is like stargazing: what we are currently observing in terms of health inequalities is partly the result of policy that was initiated years – sometimes decades – ago*” (p. 9). Research is therefore needed to understand why the social gradient in mortality has widened over the course of the twentieth century.

### Theoretical underpinning

Previous research has sought to explain the changing patterns of socio-economic inequalities, although they arrive at somewhat different answers.(12,13) First, Clouston(14) developed a four-stage framework to describe the differing relationship between socio-economic status and mortality over time, giving some consideration to why this was the case. Second, Mackenbach(4) summarized the literature on the mechanisms, defined here as causal pathways, through which socio-economic status may be related to mortality. This study also gave some consideration to how these mechanisms may have changed over time. To understand both temporal change and the mechanisms driving it, I use both approaches in this project.

First, the basis of Clouston(14) is Link & Phelan(15)’s fundamental cause theory, which was developed in response to the “risk factor epidemiology” of the late twentieth century. Individual-level, downstream factors such as diet and exercise were attributable to socio-economic differences in health. Fundamental cause theory was a push to look at factors that put people “at risks of risks,” or the more upstream factors that do not change over time, even as the more upstream risk factors might.(15) In other words, where there are socio-economic differences, so too should we expect to see socio-economic differences in health, and mortality in particular.

But socio-economic differences in mortality are not, in fact, always uniform, or indeed always present. Historical studies of the social gradient in all-cause mortality, in the Netherlands and elsewhere, often do not find consistent evidence of a social gradient.(13,16) It appears that, even in places with socio-economic differences, these differences do not result in differences in longevity. Further, studies of more contemporary populations have also found that the social gradient in specific diseases, including lung cancer and colorectal cancer, is wider today than it was in the past.(17,18)

To address such issues, Clouston(14) elaborated fundamental cause theory. The authors hypothesized that this is because socio-economic differences in mortality arise when mortality is preventable, or when interventions (either preventive or curative) were introduced. Because today, more diseases are preventable through a proliferation of different types of interventions, people enjoy longer, healthier lives.(19) This has resulted in changing and consistent patterns of socio-economic status’s relationship to (cause-specific) mortality, which can be seen over time and across diseases. The authors distilled these patterns into four stages: (1) natural mortality, in which no interventions exist, and therefore in which no socio-economic differences in mortality should be present; (2) producing inequalities, in which interventions begin to become available, and generally available first to high SES individuals – consequently, socio-economic differences in mortality begin to rise; (3) reducing inequalities, in which interventions begin to become more widely available, regardless of SES – therefore we should expect socio-economic differences begin to decrease; and (4) reduced mortality/disease death, in which the intervention becomes universal, so that there are no differences in SES in terms of mortality, and potentially no disease. Underpinning this framework is diffusion of innovations theory, in which an innovation, in this case, a health intervention, is first adopted by people with higher SES, and then spreads to the rest of society.(20) Clouston(14) helps us understand how SES’s relationship to mortality might differ over time.

However, other mechanisms may be driving changing patterns in SES and (cause-specific mortality. Mackenbach(4) considered a wider range of mechanisms by summarizing the literature on the topic, and distilling three key mechanisms. First, resources differ based on SES, with people with high SES having greater access to resources that help prevent or more effectively treat poor health. These resources may be material, such as working conditions and healthy food, or immaterial, such as health literacy and social support.(21–25) Second, non-communicable illnesses, such as type II diabetes and CVD, are partially driven by consumption behavior, and therefore socially patterned.(26) In contrast, a larger share of deaths were historically infectious disease-related, and perhaps less socially patterned.(13) It may therefore be that the benefits of the resources to which a higher social position gives access have increased.(4) Third, intergenerational mobility, or an individual’s attainment of higher levels of education and employment than their parents, may play a role. More people with low SES in childhood are later able to attain higher levels of education and employment, although this has stagnated in recent years.(27) People who remain lower SES throughout their lifetimes may be increasingly homogeneous regarding personal characteristics associated with health behavior, such as lower cognitive ability and impulse control.(28,29) These traits may be associated with poorer health. Mackenbach(4) clearly elaborates the mechanisms underpinning widening socio-economic inequities in health, but is less specific on what temporal changes would cause them.

In this project, I see Clouston et al.(14)’s stages as the historical context within which the relative importance of Mackenbach(4)’s mechanisms changes. For example, during the producing inequalities stage, when effective interventions first become available, unequal access to material and immaterial resources is likely to be the dominant driver of a social gradient in mortality of a given disease, because higher socio-economic groups adopt innovations earlier. As effective interventions diffuse more widely, during the reducing inequalities stage, this mechanism may become relatively less important. In their stead, behavioral or ‘lifestyle-related’ mechanisms may become relatively more important. Despite the two frameworks’ clear synergies, several of Mackenbach(4)’s mechanisms may not map precisely onto Clouston et al.(14)’s stages. For instance, increasing intergenerational mobility is a social process with little direct relevance to disease intervention, and its effects on disease depend both on the extent of social mobility and on the disease stage. Still, when taken together, Clouston et al.(14) and Mackenbach(4) provide a starting point to examine both how socio-economic inequities in mortality have changed over time and why.

### Research objective

This project’s objective is to understand why socio-economic inequities in mortality have widened in the Netherlands since the mid-twentieth century, by examining how the relative importance of mechanisms linking SES to mortality have changed over time.

## 2. Methods

To do so, this project takes a mixed methods approach. The project is divided into two work packages (WPs), conceptual model building (WP1) and computational modelling (WP2).

### Case study selection

To examine how the relationship between SES and cause-specific mortality changes over time, I focus on a single cause of death across different historical periods. CVD provides a particularly useful case study because it has undergone nearly all of the stages described by Clouston et al.(14), while remaining a major contributor to population mortality. As mortality from infectious diseases, particularly tuberculosis, declined, CVD emerged as the leading cause of death in the Netherlands.(30) In the mid-twentieth century, roughly half of all deaths in the Netherlands were caused by CVD.(31) However, its share declined over the twentieth century: in 2008, CVD itself was replaced as the leading cause of death by cancer mortality.(32) Over the intervening years, CVD mortality has continued to decline, with 21% of the Dutch population dying of CVD-related causes in 2020. However, the number of individuals diagnosed with CVD has increased, pointing to the success of treatment in prolonging life.(31)

There is also pronounced socio-economic patterning to CVD mortality. The 1990s saw a decrease in social differences in CVD events and mortality.(33) However, these differences appear to have later widened and remained persistent since the mid-2010s.(34) To explore the different socio-economic phases of a given disease, and different mechanisms driving it, CVD is an excellent case study. CVD mortality is therefore the key outcome of this study.

### Complex systems approach

This study adopts a complex systems approach. This approach is increasingly popular in public health, as it encourages viewing the entire context in which a health problem arises. (35,36) Taking a systems approach involves examining the many non-linear, often interacting mechanisms that drive a specific health problem, and is therefore in contrast to unidirectional statistical analysis.(37) In this project, widening socio-economic inequities in mortality are viewed as resulting from multiple, interacting mechanisms, differing in importance by both historical period, and by individual age.

When taking a complex systems approach, researchers typically employ methods that can capture interactions, feedback and changes over time.(35) These methods can be both qualitative, quantitative or a mix of both. In terms of qualitative methods, participatory model building draws on the lived experiences of diverse stakeholders to identify key mechanisms, feedback loops and other dynamics that may not be captured in existing empirical data or theory.(38,39) Involving stakeholders therefore leads to more comprehensive and realistic understandings of phenomena.(40) Further, computational modelling, including microsimulation, enables these dynamic relationships to be formalized, as computational models can account for non-linear processes, feedback loops, and reverse causality.(41) Computational models are also very useful for testing theory, as they can compare counterfactual scenarios.(42)

This project benefits from the combination of both methods. The conceptual model that stakeholders generate will be used as a basis for the computational model. Doing so has been shown to generate more valid models.(43) By combining participatory model building with microsimulation, this project will both develop and test a systems-based explanation for the widening of socio-economic inequalities in mortality in the Netherlands.

### Patient and public involvement

Patients and members of the public will contribute to this study through the participatory sessions. Individuals with lived experience of poverty and CVD, alongside representatives from patient organizations and other stakeholders, will contribute to the development and refinement of the conceptual model. Their input will directly inform the structure of the microsimulation model and the co-design of a layman’s summary of findings.

### Qualitative conceptual model building (WP1)

This project begins by building a conceptual model, or an abstracted, simplified diagram, to understand how different factors relate to one another. The conceptual model explains widening socio-economic inequities in CVD in the Netherlands. To do so, this project uses Group Model Building methodology, which was developed to leverage participatory input in conceptual models.(38) This methodology was also developed to capture complex system dynamics, such as non-linearities, where changes in exposure do not produce proportional changes in an outcome, feedback loops, where changes in one factor ultimately influence the same factor through a chain of events, and delays, or time lags between a change in one part of a system and its effects becoming apparent elsewhere in the system. The conceptual model potentially including such system dynamics will be developed over three sessions alongside stakeholders.

Ten to fifteen stakeholders will be recruited for each of these sessions, in line with recommendations for conceptual model building.(39) This number of participants has been found to elicit a variety of perspectives, while still allowing participants space to speak in sessions.(44) Participants will be recruited purposively to reflect a diversity of perspectives on SES and CVD. Participants may include: policymakers, members of patient and doctor organizations, members of charities aimed at tackling CVD, people with lived experience of poverty, and municipal health workers. Potential participants will initially be approached by the projects principal investigator (KT) via email. To ensure continuity across sessions, the same participants will ideally participate in all three sessions, although participant drop-outs may necessitate recruiting new participants.(38)

The three sessions will be held in-person. Sessions will last between two and four hours each, with each workshop scheduled roughly six weeks apart. Along with the participants, at each session, there will be a facilitator leading the discussion, a recorder taking notes, and a modeler constructing the conceptual model in real time. The modeler will use Group Model Building software, such as Vensim, to do so.(45) The evolving model will be projected to participants on a large screen, allowing iterative refinement throughout the sessions. All sessions will be audio-recorded and transcribed verbatim. Field notes taken by the recorder and successive versions of the conceptual model will also be retained as data sources.

The structure of the sessions will be adapted from validated group model building scripts.(46) In session one, participants will define the boundaries of the system and identify the key factors that influence socio-economic inequalities in CVD in the present day. For instance, smoking might be identified as one such factor. Through facilitated discussion, they will develop a shared understanding of the problem and generate an initial overview of the factors and actors involved.

In the second session, participants will connect the identified variables into a causal structure by specifying the direction of relationships between them (for example, whether smoking increases CVD risk, or whether developing CVD subsequently influences smoking behavior). Participants will also identify systems dynamics, such as non-linearities. For instance, participants might suggest that the relationship between smoking and CVD risk is non-linear: relatively low levels of cigarette consumption can already produce a disproportionately large increase in cardiovascular risk.(47)

In the third session, participants will use the present-day conceptual model as a starting point to consider how this system they have visualized has changed over time. They will identify which mechanisms have strengthened, weakened, emerged, or disappeared since the mid-twentieth century. Participants will do so by considering developments such as advances in medical treatment, changes in the food environment, and broader social and economic changes. For instance, changing smoking rates and/or changing policies around tobacco consumption may be included. Also in this session, a summary of Mackenbach(3)’s theory of the emergence and persistence of socio-economic differences in health will be introduced as a theoretical point of comparison. Participants will compare their conceptual model with this framework, identifying areas of agreement, disagreement, and additional mechanisms not captured by the existing theory. These discussions will inform the microsimulation by identifying which mechanisms have changed over time, including changes in their relative importance or strength.

To recognize stakeholders’ contributions, session three also involves co-designing a product showcasing the results of the conceptual model in an accessible format. This could take the form of, e.g. an infographic, and/or an executive summary of key findings.

Following each workshop, the research team will review the draft conceptual models, workshop recordings, and field notes to clarify terminology, remove duplicate variables, and prepare an updated conceptual model for the subsequent workshop. Changes made between workshops will be documented.

The final output of WP1 will be a stakeholder-derived conceptual model of socio-economic inequalities in CVD, with a particular view to how these dynamics have changed since the mid-1950s. The workshop findings will then guide how these historical changes are represented and quantified in the computational model. This process has been shown to increase the face validity of computational models.(48)

### Computational modelling (WP2)

In WP2, the co-created conceptual model will provide the foundation for a microsimulation, a computational model that will simulate the life courses of individuals within the broader Dutch social system to investigate the widening of socio-economic health inequities. Microsimulations are especially well-suited to representing how structural determinants, such as socio-economic status (SES), shape individual trajectories, for instance health across the life-course, and how these individual trajectories aggregate to produce population-level patterns.(49,50) Like group model building, microsimulations can incorporate system dynamics.(51) By translating the theoretical relationships identified in the conceptual model in WP1 into a computational one, WP2 enables us to quantify the contribution of different mechanisms for widening health inequities over time.(42,52)

The microsimulation will be built in Python, using the conceptual model developed in WP1 to determine which characteristics and mechanisms are represented and how they are related. These individual characteristics, or state variables, may be time-invariant (remaining fixed throughout the simulation, such as sex) or time-varying (changing over the life course, such as age, SES, or smoking behavior). In yearly time steps, individuals may experience transitions that change time-varying state variables, such as changes in SES or smoking behavior, as well as health transitions, including developing CVD and death. The probability of each transition will depend on an individual’s current states and, where relevant, their previous state variables and transitions. Mechanisms identified in the conceptual model will be represented through these dependencies. For example, a mechanism linking SES to smoking and smoking to CVD would be represented by SES influencing the probability of smoking and smoking, in turn, influencing the probability of developing CVD. In this way, the model represents how individual characteristics and experiences accumulate and interact over the life course to influence CVD risk and mortality.

The microsimulation will comprise a single computational model with a common structural framework applied to successive twenty-year historical periods from 1950 onward. For each period, a separate synthetic population will be generated to represent the Dutch population at the beginning of that period in terms of demographic, socio-economic and health characteristics relevant to CVD, and will be simulated over the subsequent 20 years. The state variables and mechanisms included in the model will remain constant across periods, as defined by the conceptual model developed in WP1. However, the strength of individual mechanisms may vary across periods, reflecting historical changes in the processes underlying socio-economic health inequities. The value of a state variable or mechanism may also be set to zero, effectively silencing the corresponding mechanism if it was not relevant to a particular period. Both the synthetic populations and the microsimulation will be specified separately for each twenty-year period, using longitudinal data from the Historical Sample of the Netherlands (HSN), the System of Social-statistical Datasets (SSD), and linked datasets.(53–55)

Before using the microsimulation to investigate explanations for widening socio-economic health inequities, the model will first be validated against observed historical trends. In twenty-year periods, the model will be run using the estimated parameter values, and the resulting socio-economic inequalities in CVD and mortality will be compared with those observed in a hold-out sample not used for parameter estimation. This will assess whether the model can reproduce historical patterns of socio-economic inequalities in CVD and mortality before it is used for subsequent simulation experiments.

Second, counterfactual simulation experiments will be used to estimate the contribution of individual mechanisms to the widening of socio-economic inequities in CVD mortality. Beginning with the parameter values for an earlier historical period, the parameters associated with one mechanism, such as smoking, will be replaced with those estimated for a later period, while all other parameters remain fixed at their earlier-period values. The resulting change in SES differences in CVD mortality will indicate how much the historical change in that mechanism contributes to the widening of socio-economic inequities in CVD mortality. This process will be repeated for each mechanism. Parameters associated with multiple mechanisms will then be replaced simultaneously to assess their joint and interacting contributions. For example, updating smoking parameters alone may explain 20% of the widening and updating diet parameters alone 15%, whereas updating both together may explain 50%, indicating that the two mechanisms reinforce one another. These experiments will identify the relative importance of different mechanisms, individually and in combination, to the widening socio-economic gradient in CVD mortality in the Netherlands.

## Ethics and dissemination

This project was approved by the Social Sciences Research Ethics Committee of Wageningen University (approval number 2026-203). All participants in the Group Model Building workshops will receive written information about the study and provide informed consent prior to participation. Participation will be voluntary, and participants may withdraw at any time without providing a reason. Participants will receive €40 per workshop attended to compensate their time, in addition to reimbursement of reasonable travel expenses and refreshments during the workshops.

Only data necessary to address the research objectives will be collected. Personal identifiers and other sensitive information will be stored separately from research data, with access restricted to the research team. Data will be anonymized prior to analysis wherever possible, raw identifiable data will be securely deleted at the end of the project, and anonymized research data will be archived for a minimum of ten years in accordance with institutional policy.

The microsimulation code and accompanying documentation will be openly available via GitHub. Due to data-sharing agreements with the providers, the underlying datasets cannot be made publicly available. Project findings will be disseminated through peer-reviewed open-access publications, conference presentations, and a public summary of findings.

## 3. Discussion

This protocol details a mixed-methods study to investigate why socio-economic inequities in CVD have widened in the Netherlands since the mid-twentieth century. The project will first leverage stakeholder knowledge to develop a conceptual model of the mechanisms underpinning widening socio-economic inequities in CVD. The resulting microsimulation will enable both historical comparisons and counterfactual simulation experiments, to quantify the contribution of competing mechanisms to widening inequalities.

This project has a number of strengths. First, this project integrates qualitative and quantitative methods. Rather than relying solely on existing theory, the conceptual model will be co-created with stakeholders and translated directly into the microsimulation, increasing the face validity and policy relevance of the simulation.(43) Further, although long-run dynamic microsimulations have previously been developed in fields such as economics and demography, this approach has rarely been applied to understanding the historical development of socio-economic health inequities.(56,57)

The project also has several limitations. First, while rich contemporary linked data are available to parameterize many mechanisms, historical data are more limited and some parameters may need to be estimated using proxy measures or assumptions. Second, discrepancies may arise between mechanisms identified through participatory sessions and those that have been identified as important in the literature. However, these discrepancies may identify important evidence gaps and priorities for future research. Even with these limitations, this project takes a rigorous approach to investigate the complex mechanisms underlying widening socio-economic inequities in health.

## Data Availability

This paper is a study protocol, and does not use any data.

## Author Statement

KT – Conceptualization; Methodology; Writing – original draft; Writing – review and editing.

## Data statement

No data were collected in the drafting of this protocol paper.

